# Genomic Epidemiology of Methicillin-Resistant *Staphylococcus aureus* in Two Cohorts of High-Risk Military Trainees

**DOI:** 10.1101/19011445

**Authors:** Robyn S. Lee, Eugene V. Millar, Alanna Callendrello, Caroline E. English, Emad M. Elassal, Michael W. Ellis, Jason W. Bennett, William P. Hanage

**Affiliations:** Epidemiology Division, Dalla Lana School of Public Health, University of Toronto, Toronto, Canada; The Center for Communicable Disease Dynamics, Department of Epidemiology, Harvard T.H. Chan School of Public Health, Boston, MA, USA; Infectious Disease Clinical Research Program, Department of Preventive Medicine and Biostatistics, Uniformed Services University of the Health Sciences, Bethesda, MD, USA; Henry M. Jackson Foundation for the Advancement of Military Medicine, Bethesda, MD, USA; University of Toledo College of Medicine and Life Sciences, Toledo, OH, USA; Walter Reed Army Institute of Research, Silver Spring, MD, USA; Department of Medicine, Uniformed Services University of the Health Sciences, Bethesda, MD, USA

**Keywords:** Methicillin-resistant Staphylococcus aureus, genomic epidemiology, military, community-acquired MRSA, transmission

## Abstract

**Background:** MRSA skin and soft tissue infection (SSTI) is a significant cause of morbidity in military trainees. To guide interventions, it is critical we understand the epidemiology of MRSA in this population.

**Methods:** Two cohorts (‘companies’) of US Army Infantry trainees (N=343) at Fort Benning, GA, USA, were followed during their training cycles (Jun.-Dec. 2015). Trainees had nares, oropharynx, perianal and inguinal areas swabbed for MRSA colonization at five ∼2-4 week intervals, and monitored for SSTI throughout training. Epidemiological data were collected. Isolates were sequenced using Illumina HiSeq and NovaSeq. Single-nucleotide polymorphisms and clusters were identified. Multi-locus sequence type (MLST) and antimicrobial resistance genes were predicted from *de novo* assemblies.

**Results:** 87 trainees were positive at least once for MRSA (12 had SSTI, 2 without any colonization). Excluding those positive at baseline, 43.7% were colonized within the first month of training. 244/254 samples were successfully sequenced (including all SSTI). ST8 (n=135, 100% of SSTI), ST5 (n=81) and ST87 (n=21) were the most represented. Three main Clusters were identified, largely corresponding to these STs. Sub-analyses within Clusters showed multiple importations of MRSA, with transmission subsequently predominantly within, rather than between, platoons in each company. Over 50% of trainees were colonized only at other anatomical sites; restricting analyses to nares missed substantial transmission.

**Conclusions:** Serial importations of MRSA into this high-risk setting likely contribute to the ongoing burden of MRSA colonization and infection among military trainees. Sampling multiple anatomical sites is critical for comprehensive characterization of MRSA transmission

**Summary:** US Infantry trainees were followed through training for MRSA skin and soft tissue infection, swabbing for colonization at 2-4 week intervals. Sequencing suggests serial importations of diverse strains on base, followed by transmission mostly within platoons, involving multiple anatomical sites.

## Introduction

Community-associated methicillin-resistant *Staphylococcus aureus* (CA-MRSA) is a major cause of skin and soft tissue infections (SSTI) [1]. Military personnel have an elevated risk of MRSA SSTI compared to the general population with new recruits disproportionately affected [2, 3]. SSTI outbreaks compromise not only training cycles, but also operational readiness among deployed forces [4]. The annual cost of SSTIs to the US military is approximately 14-32 million US dollars [5]. Past SSTI prevention strategies in military populations have included nasal decolonization [6] as well as chlorhexidine-based body wash [7, 8], based on the principle that MRSA colonization precedes and is necessary for infection [9]. However, neither strategy prevented SSTI in these high-risk populations [6, 8].

To develop effective prevention strategies, it is critical we understand the epidemiology and transmission dynamics of MRSA in this population. Recently, whole genome sequencing (WGS) has been used to describe the epidemiology of MRSA colonization [4] and infection among US Army trainees [10]. A key limitation of this study was that WGS was restricted to trainees with pulse-field gel electrophoresis (PFGE) type USA300 MRSA, the most common cause of CA-MRSA SSTI. As such, the conclusions that could be drawn about overall MRSA transmission dynamics, and prevalence of different strains circulating in this population, were greatly limited. To address this concern, we conducted a prospective cohort study of MRSA in the same trainee population, following them throughout their training cycle for colonization and infection events. All MRSA-positive samples from multiple anatomical sites were included.

Our study found a high prevalence of MRSA colonization at baseline, comprising a genetically diverse population imported from across the US by recruits entering basic training. This was followed by substantial person-to-person transmission, particularly among soldiers belonging to the same ‘platoons’ (i.e., those in the same training company who share the same living quarters). Moreover, the observed transmission events frequently involved different anatomical sites - irrespective of nasal colonization. As nasal swabs are routinely and almost exclusively used to detect MRSA colonization in clinical settings, this suggests MRSA transmission may be drastically under-estimated with current infection control methodology – a finding with significant implications for the control of MRSA.

## Methods

### Overview

We conducted a longitudinal cohort study of SSTI among US Army Infantry trainees at Fort Benning, Georgia, from June-December 2015. Two companies (designated A and B) were included, which commenced the 14-week training cycle at two different timepoints during peak SSTI seasons (June and September, respectively). Infantry training companies are comprised of ∼200 soldiers, further segregated into four platoons comprised of ∼50 soldiers. See **Supplementary Methods** for additional detail.

### MRSA screening

MRSA colonization swabs were obtained at baseline and at ∼2-4 week intervals over five study visits. [6] Anterior nasal, oropharyngeal, inguinal and perianal swabs were collected (see **Supplementary Methods**). Swabs were initially processed at the Benning Martin Army Community Hospital clinical microbiology laboratory as previously described [8].

### Epidemiologic data collection

Trainees completed questionnaires at baseline for demographic and MRSA risk factor information, and at follow-up visits to collect data on personal hygiene practices.

### SSTI case definition

Trainees were considered to have probable SSTI if they presented with cellulitis, abscess, folliculitis, impetigo, paronychia, infected blister, or pilonidal cyst; SSTI cases were defined as those with *S. aureus* cultured from the clinical site. Case detection is described in the **Supplementary Methods**.

### Genome sequencing and bioinformatics

See **Supplementary Methods** for details. In brief, genomes were sequenced using the Illumina HiSeq and NovaSeq platforms. As most samples were known to be USA300, we initially aligned reads to the USA300 TCH1516 reference genome (National Center for Biotechnology Information [NCBI] Accession CP000730.1) using the Burrows-Wheeler Aligner MEM algorithm (v.0.7.15, [12]) and identified variants compared to this reference. Prophage sequences were identified using Phaster (http://phaster.ca, [13]) and SNPs in these regions were masked.

Concatenated SNP alignments were used to generate maximum likelihood trees with IQ-TREE (v.1.6.8, [14]). To infer a more accurate phylogeny for the analysis including all STs, we masked predicted recombination sites using Gubbins (v.2.3.4, [15]). Hierarchical Bayesian Analysis of Population Structure (RhierBAPS v.1.1.0, [16]) was used to discriminate clusters of closely-related samples in R (v.3.5.2). Analyses were then repeated for each major Cluster identified using a closely-related reference genome.

*De novo* assemblies were also generated, using SPAdes (v.3.8.0, [17]) and Unicycler (v.0.4.7, Wick R, https://github.com/rrwick/Unicycler). Sequence Types (STs) according to the multi-locus sequence typing (MLST) scheme [18] were predicted using ‘mlst’ (v.2.10, Seemann T, https://github.com/tseemann/mlst) and Abricate (v.0.8, Seemann T, https://github.com/tseemann/abricate) was used with ResFinder [19] and the Virulence Factor DataBase (VFDB, [20]) to detect genes associated with antimicrobial resistance and virulence, respectively. *mupA* and *qacA* genes were predicted using gene-puller.py (https://github.com/kwongj/gene-puller).

### Statistical analysis

Categorical comparisons were done using the Chi-Square test where cell counts > 5, or Fisher’s Exact test if otherwise. Distributions were compared using the Wilcoxon-Mann-Whitney test. Conditional logistic regression was used to compare hygiene variables between trainees who became colonized with time-matched, MRSA-negative controls (**Supplementary Methods)**. Analyses were done in Stata15 (StataCorp, College Station, TX, USA).

## Data availability

Sequence data is on NCBI’s Sequence Read Archive under BioProject PRJNA587530.

## Ethics approval

The study was approved by the Uniformed Services University Institutional Review Board (IDCRP-090) and Harvard T.H. Chan School of Public Health Institutional Review Board (IRB17-0802).

## Results

We followed two Infantry training companies in the summer and fall of 2015. 343 trainees participated (132 and 211 in companies A and B, respectively), with 86.6% (297/343) completed all study visits (**Supplementary Results**). 87 (25.4%) of trainees were MRSA-positive during the study; 85 had at least one positive colonization swab and 12 were diagnosed with MRSA SSTI. Among the latter, only seven were colonized prior to or at the same time as their SSTI (five with the infecting strain); three were colonized only at later timepoints, and two were never colonized (**Supplementary Table 1**).

**Table 1.** Clinical and epidemiological characteristics of trainees at baseline (N=343)

| Characteristic | Colonized and/or SSTI (n=87) | Never colonized and no SSTI (n=256) | P value |
| --- | --- | --- | --- |
| Age, average (SD) | 19.2 (1.7) | 19.4 (2.5) | 0.42<sup>a</sup> |
| Hispanic ethnicity, n (%) | 15 (17.2) | 40 (15.6) | 0.72<sup>b</sup> |
| Race, n (%) |  |  |  |
| White | 64 (73.6) | 182 (71.1) |  |
| Black | 8 (9.2) | 23 (9.0) |  |
| Asian | 0 (0) | 6 (2.3) |  |
| American Indian | 1 (1.1) | 5 (2.0) |  |
| Pacific Islander | 0 (0) | 3 (1.2) |  |
| Other<sup>c</sup> | 7 (8.0) | 20 (7.8) |  |
| > 1 race reported | 7 (8.0) | 17 (6.6) | 0.88<sup>d</sup> |
| Lived alone prior to training, n (%) | 5 (5.7) | 10 (3.9) | 0.54<sup>d</sup> |
| Lived with pets/livestock, n (%) | 64 (73.6) | 177 (69.1) | 0.44<sup>b</sup> |
| Antibiotics in previous 6 months, n (%) | 6 (6.9)<sup>e</sup> | 16 (6.3) | 0.83<sup>b</sup> |
| Admitted to hospital in past year, n (%) | 1 (1.1) | 6 (2.3) | 0.68<sup>d</sup> |
| Worked in a hospital in past year, n (%) | 0 (0) | 8 (3.1) | 1.0<sup>d</sup> |
| Lived with someone who was hospitalized/worked in hospital in past year, n (%) | 1 (1.1) | 69 (27.0) | 0.47<sup>d</sup> |
| Known/suspected SSTI/MRSA infection in past year, n (%) | 2 (2.3) | 2 (0.8) | 0.27<sup>b</sup> |
| Prior contact with MRSA SSTI in 3 months prior to start of training, n (%) | 1 (1.1) | 4 (1.6) | 1.00<sup>b</sup> |

Note, all trainees on base were male during the study year. SSTI = skin and soft tissue infection ; MRSA = methicillin-resistant *S. aureus*. <sup>a</sup> T-test with unequal standard deviations. <sup>b</sup> Chi-square test with 1 df. <sup>c</sup> Includes 28 individuals who self-reported as “Hispanic” and selected race “Other”. <sup>d</sup> Fisher’s Exact test. <sup>e</sup> One individual with prior antibiotic exposure had an SSTI; none of those with SSTI had hospitalization, prior SSTI themselves or contact with someone with SSTI in the three months prior to arrival to Fort Benning.

Among those colonized, thirty-five had a single positive sample; the oropharynx and perianal sites were most frequently colonized, each representing 31.4% of swabs, followed by nasal (20.0%), and inguinal (17.1%) sites. Fifty trainees were colonized at multiple anatomical sites and/or timepoints. The number of repeat positive samples did not differ significantly by anatomical site of isolation (p = 0.16), although inguinal swabs tended to be less frequently positive (**Figure 1**).

**Figure 1.**
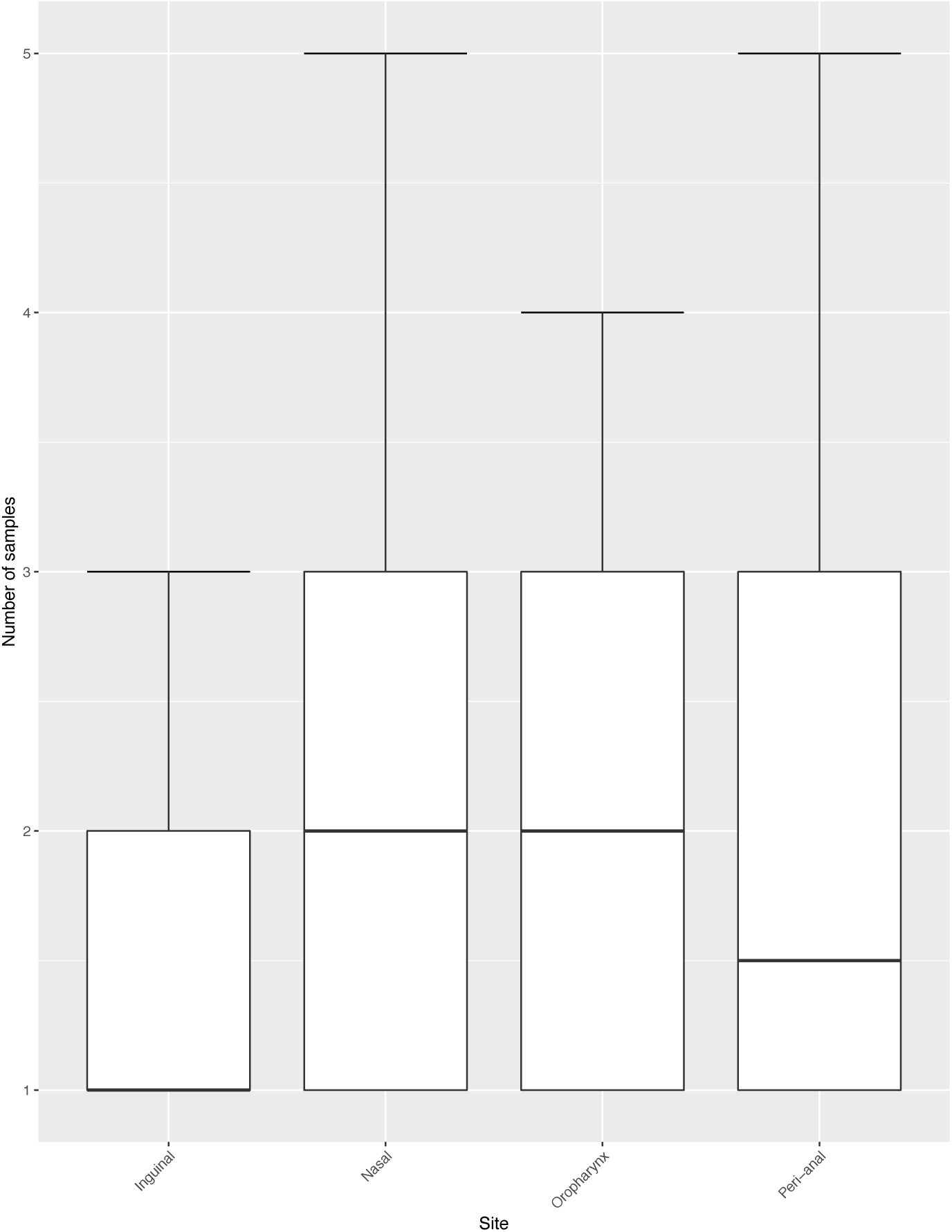
Number of isolates per trainee, by anatomical site of isolation. 85 trainees had at least one non-clinical sample positive for MRSA during the study. Includes samples that were not available/sequenced.

### Risk factors for colonization or infection

There were no significant differences in age, self-reported race, or MRSA risk factors at baseline (**Table 1**) comparing those with colonization/infection to those who were never MRSA-positive. Epidemiological characteristics of SSTI cases are described in the **Supplementary Results**.

### Timing of MRSA acquisition

Twenty-seven trainees were colonized at baseline (see **Supplementary Results**). Excluding these, most trainees appeared to have acquired MRSA shortly after starting training (**Figure 2**); 43.7% of all positive samples were collected during the first month of training. There were no significant differences in personal hygiene practices over the same timepoint between trainees with new colonization and those who were never positive (**Supplementary Results, Supplementary Table 2**).

**Table 2.** Genomic characteristics of included isolates (N=244)

| *In silico* prediction | Colonization, n=232 | Clinical, n=12 |
| --- | --- | --- |
| MLST, n (%) |  |  |
| ST8 | 123 (53.0) | 12 (100) |
| ST5 | 81 <sup>a</sup> (34.9) | 0 (0) |
| ST87 | 21 (9.1) | 0 (0) |
| Other STs <sup>b</sup> | 4 (1.7) | 0 (0) |
| SLV of ST8 | 1 (0.4) | 0 (0) |
| Novel STs <sup>c</sup> | 2 (0.9) | 0 (0) |
| lukS/lukF-positive, n (%) | 108 (46.6) | 12 (100) |

<sup>a</sup> ‘MLST’ refers to multi-locus sequence type, while ‘SLV’ refers to a single locus variant, wherein one allele differs from the listed ST. Three colonization samples were PCR-positive for *mecA* but negative *in silico*: 1133\_2P, 1134\_5N, 2069\_2I. <sup>a</sup> This includes isolate 2014\_3P which has two alleles of the *tpi* gene (1,99). <sup>b</sup> This includes ST15, ST30, ST45, ST72. <sup>c</sup> Includes a triple locus variant of ST8.

**Figure 2.**
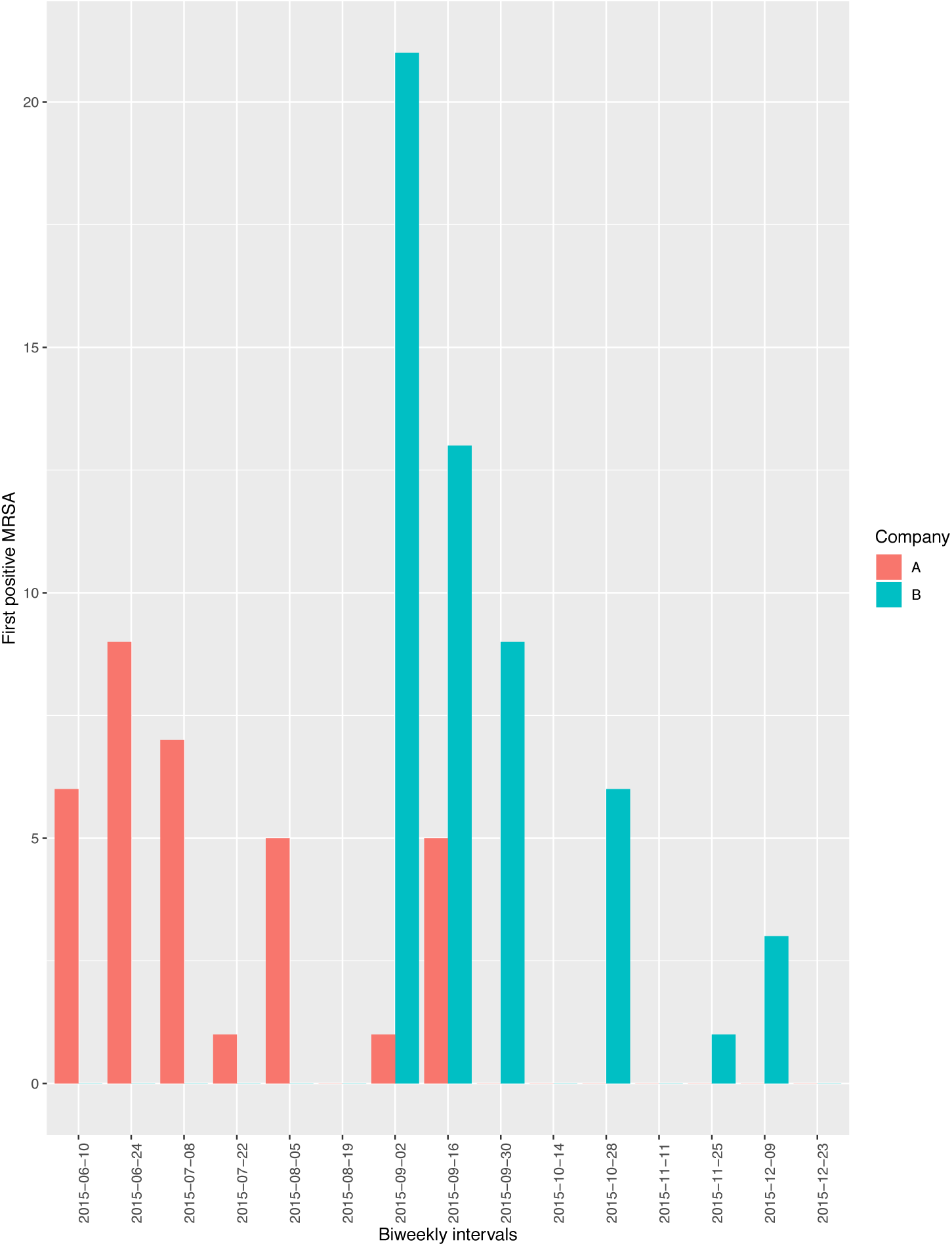
Timing of first MRSA positive samples collected from each trainee, by company at Fort Benning, Georgia, 2015. In total, 87 trainees were positive for MRSA from the two Companies under study. 27 trainees were positive at baseline; these samples were collected within 1-2 days of arrival to base, while all trainees were at one central location.

### Genomic analyses

High-quality sequencing data was available for 244/254 (96.1%) MRSA-positive samples from 85 individuals; further analyses presented are restricted to this subset. ST8 (USA300) was the dominant strain identified (**Table 2**), and included all cases of SSTI. The virulence factor Panton-Valentine Leukocidin was present in all clinical isolates (as demonstrated by intact *lukS* and *lukF* genes) and nearly half of colonization isolates (**Table 2**). All samples were PCR-positive for *mecA*, while 98.8% were predicted to have *mecA in silico*; 2/3 isolates lacking *mecA in silico* were different STs than other isolates from those trainees, indicating that mixed genotypes may have been present at sampling (**Supplementary Table 3**). To assess the potential utility of mupirocin and chlorhexidine for decolonization, *mupA* and *qacA* genes were predicted. *mupA* was only detected in colonization isolates (all ST5) from four trainees (and one ST8 clinical isolate from another trainee), while *qacA* was detected in colonization isolates from three trainees. Correlation between PFGE and ST are shown in **Supplementary Table 4**.

**Table 3.** Pairwise core single nucleotide polymorphism (SNP) distances within sub-groups of Clusters 1, 2 and 4

| Cluster | Sub-group | Number of isolates | Number of trainees | SNP distance between trainees, median (range) | SNP distance within trainees, median (range) | Number of trainees with positive at baseline, n (%) | Number of trainees with nasal samples, n (%) | Number of trainees with clinical samples, n (%) |
| --- | --- | --- | --- | --- | --- | --- | --- | --- |
| 1 | 1-1 | 69 | 24 | 7 (0-383) | 0 (0-9) | 4 (16.7) | 12 (50.0) | 6 (25.0) |
|  | 1-2 | 16 | 8 | 4 (0-9) | 0 (0-4) | 0 (0) | 3 (37.5) | - |
|  | 1-3 | 7 | 2 | 2 (1-3) | 0 (0-1) | 1 (50.0) | 1 (50.0) | - |
|  | 1-4 | 11 | 5 | 1 (0-3) | 0 (0-2) | 0 (0) | 2 (40.0) | 3 (60.0) |
|  | 1-5 | 4 | 2 | 159 (157-161) | 3 (2-4) | 1 (50.0) | 0 (0) | - |
|  | 1-6 | 8 | 3 | 21 (0-22) | 1 (0-7) | 1 (33.3) | 1 (33.3) | 2 (66.7) |
|  | 1-7 | 14 | 2 | 0 (0-8) | 1 (0-10) | 1 (50.0) | 1 (50.0) | 1 (50.0) |
|  | 1-8 | 6 | 3 | 0 (0-2) | 1.5 (0-3) | 3 (100) | 0 (0) | - |
|  | 1-9 | 4 | 1 | - | 8.5 (2-15) | 1 (100) | 0 (0) | - |
| 2 | 2-1 | 9 | 5 | 4 (1-116) | 0 (0-3) | 1 (100) | 2 (40.0) | - |
|  | 2-2 | 51 | 23 | 1 (0-655) | 1 (0-3) | 3 (13.0) | 7 (30.4) | - |
|  | 2-3 | 11 | 2 | 0 (0-4) | 1 (0-5) | 1 (50.0) | 1 (50.0) | - |
|  | 2-4 | 3 | 1 | - | 1 (0-1) | 1 (100) | 0 (0) | - |
|  | 2-5 | 2 | 1 | - | 0 (0-0) | 1 (100) | 0 (0) | - |
|  | 2-6 | 5 | 1 | - | 0 (0-0) | 1 (100) | 0 (0) | - |
| 4 | 4-1 | 12 | 6 | 0 (0-2) | 1 (0-3) | 6 (100) | 1 (16.7) | - |
|  | 4-2 | 9 | 5 | 0 (0-0) | 0 (0-0) | 0 (0) | 4 (80.0) | - |

Clusters were identified in the analysis including all samples, using hierBAPS [16]. Core SNPs within each Cluster were then identified by repeating the analysis, but aligning the reads from isolates within each Cluster to a closely-related reference from the most represented sequence type in that Cluster. hierBAPS [16] was then run again for each Cluster using these new alignments as input (to obtain the highest resolution), and sub-groups of closely-related isolates were identified within each Cluster. The number of core SNPs were tabulated within each sub-group and are comparable within, but not across, Clusters (as each Cluster has a different core genome). Cluster 1 sub-groups were separated by a minimum of 34 core SNPs, Cluster 2 sub-groups were separated by a minimum of 150 core SNPs, and Cluster 4 sub-groups were separated by a minimum of 100 core SNPs. Cluster 1 was predominantly ST8 (n=136/139); three non-ST8 isolates were included based on hierBAPS analysis, including 1133\_2P (a single locus variant of ST8), 1105\_5P (a triple locus variant of ST8) and 1133\_2P (ST72, a triple locus variant of ST8). These three non-ST8 isolates were included in sub-group 1-1, and were a maximum of 383 core SNPs to another other sample within this sub-group. Cluster 2 was comprised of all ST5 isolates, while Cluster 4 was comprised of all ST87 isolates.

### Strain replacement

Strain replacement was rare (**Supplementary Figure 1**), occurring only four times during the study (excluding co-colonization). Two instances involved different STs; trainee 2013 was initially colonized with an ST5 strain but developed an ST8-SSTI, while 2031 was colonized with ST8 but later colonized with ST5. The remaining instances involved trainees 1007 and 2061. These trainees had isolates from the same ST that were separated from their later isolates by > 97 core SNPs, suggestive of exogenous re-infection.

### Overall transmission analyses

See **Supplementary Results** for quality metrics. The recombination-adjusted maximum likelihood tree is shown in **Figure 3**, excluding one highly divergent isolate (1134_5N, see **Supplementary Results**). Five Clusters were identified, with three containing >1 isolate; Cluster-1 (n=139, 97.1% ST8); Cluster-2 (n=81, all ST5); and Cluster-4 (n=21, all ST87). Pairwise adjusted core SNP distances between isolates indicate that transmission was predominantly within, rather than between, platoons in the same Companies (**Figure 4**).

**Figure 3.**
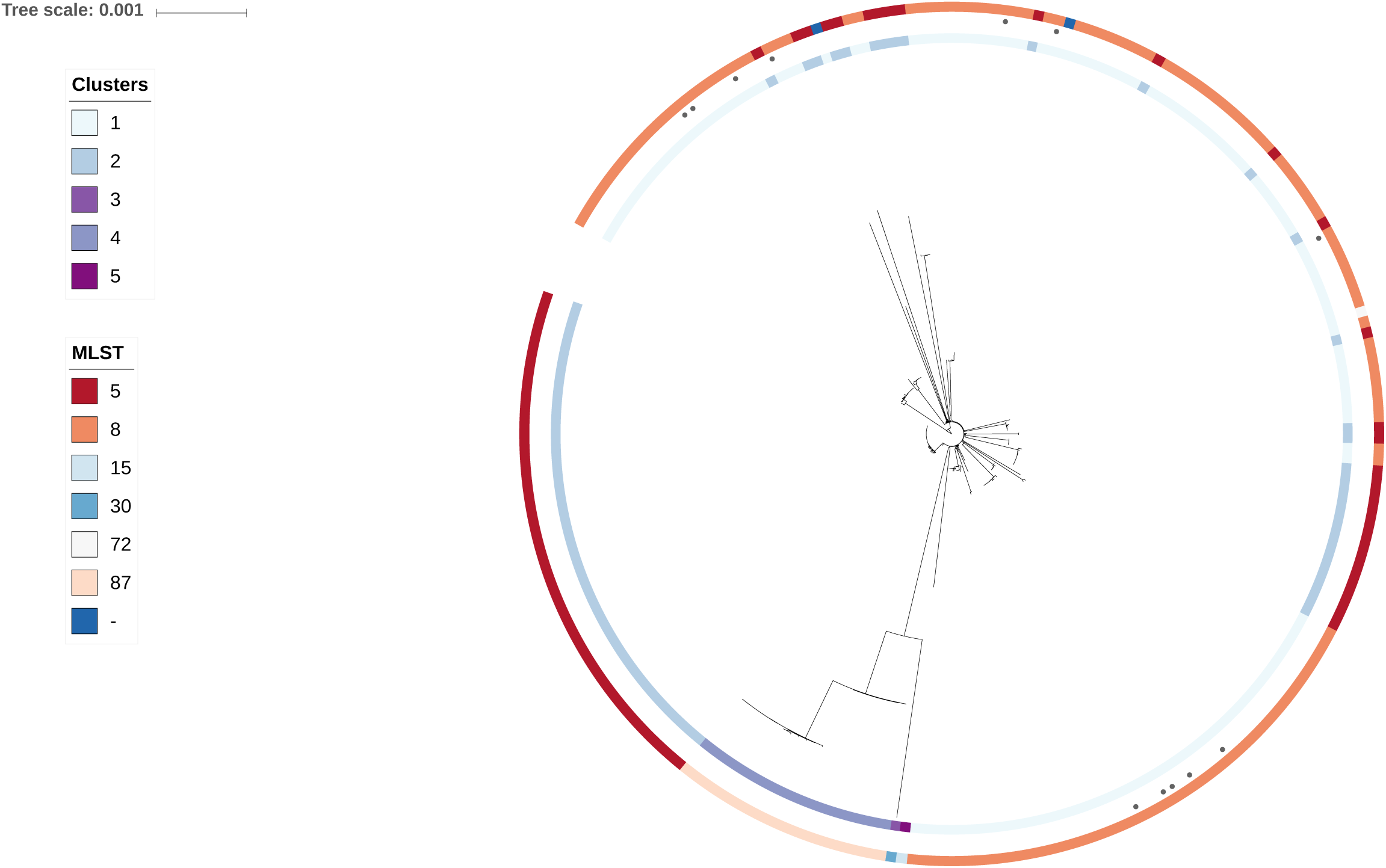
Maximum likelihood tree for colonization and clinical samples from trainees undergoing basic training at Fort Benning, Georgia in 2015. One isolate (1134_5N) was highly divergent from all others, at > 18,500 core SNPs compared to < 1000 SNPs for the next most divergent. Therefore, this isolate was excluded from this analysis. A concatenated SNP alignment of the remaining isolates was produced using snp-sites [28] and IQ-Tree [14] (v1.6.8) was used to produce the maximum likelihood tree, using the TVMe model with ascertainment bias correction. Model selection was based on the lowest Bayesian Information Criterion. Hierarchical Bayesian Analysis of Population Structure (hierBAPS) [16, 29] was used to identify clusters with the full alignment as input. The tree was visualized using the Interactive Tree of Life [30]. Samples from skin and soft tissue infection are indicated.

**Figure 4.**
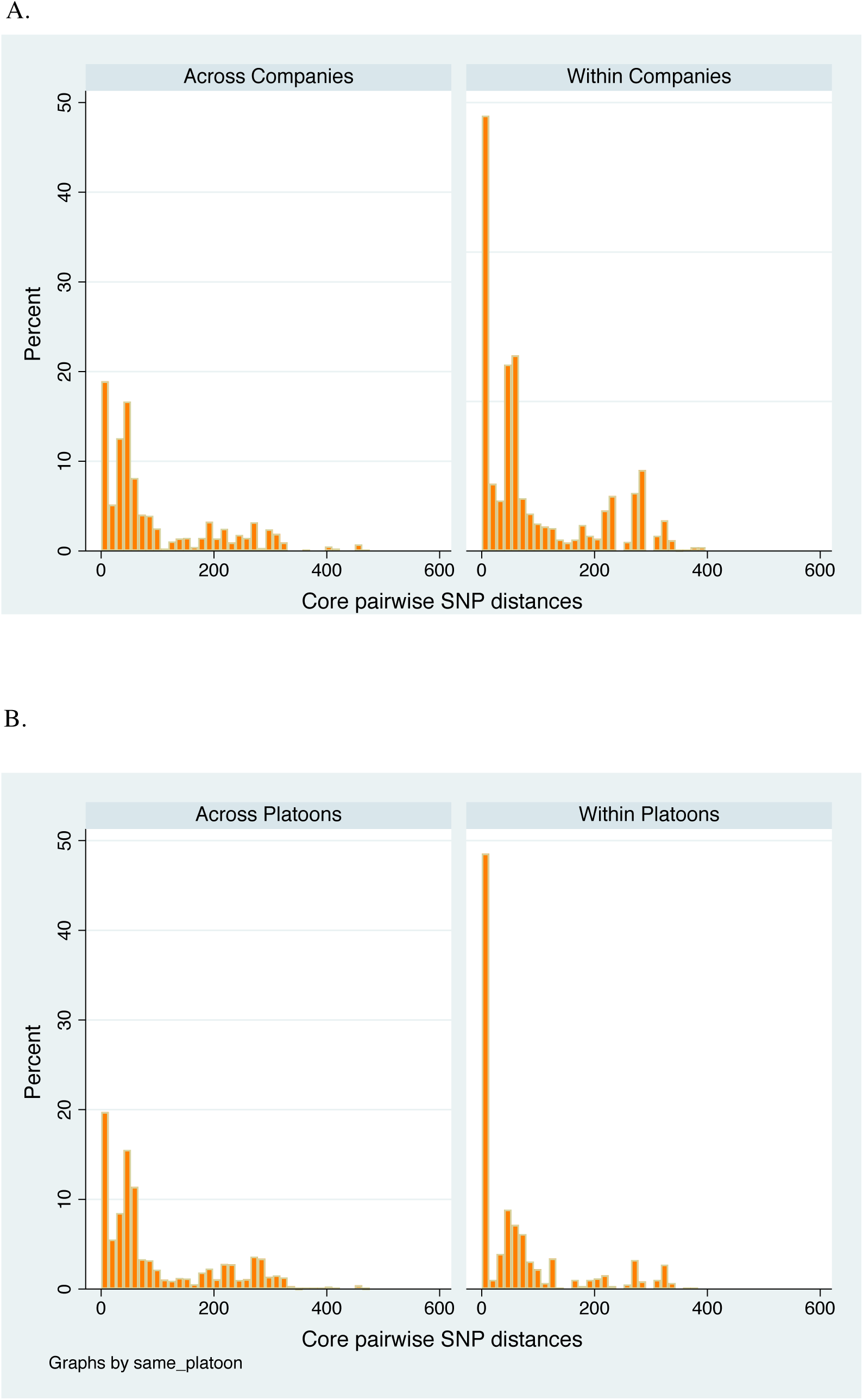
Recombination-adjusted core pairwise SNP distributions by company and platoon. **A. Pairwise core SNPs across and within companies.** **B. Pairwise core SNPs across and within platoons.** Core SNPs were identified compared to the USA300 reference (excluding isolate 1134_5N due to its high divergence from all others isolates in the dataset). This resulted in 29,403 pairwise comparisons between trainees, including repeat samples from the same individuals. Retaining closely-related samples from the same trainees would be expected to artificially reduce the pairwise distances within companies and platoons. Therefore, where repeat samples from a single participant were within 5 SNPs of one another, only one of these was retained for this analysis. This threshold was chosen because it represented 90% of pairs from the same trainees. In total, 3,739 pairwise comparisons were therefore included in this analysis. Note that all trainees in the same platoon are, by default, in the same company.

### Sub-analyses of transmission, by Cluster

To provide the highest resolution, analyses were re-run for each Cluster using a closely-related reference (**Supplementary Table 5**). As ST8s represent the largest proportion of samples collected, the maximum likelihood tree for Cluster 1 is shown in **Figure 5** (see **Supplementary Figures 2** and **3** for Clusters 2 and 4, respectively). Within Cluster 1, there were nine sub-groups each representing isolates from 1 to 24 trainees. These sub-groups were separated from one another by a minimum of 34 SNPs (**Table 3**). In nearly all sub-groups, there were isolates from different trainees that shared close genetic relationships, indicative of recent transmission (**Table 3**). Overlaying company and platoon (**Figure 5**) suggests that such transmission was largely restricted to the same platoons, with a small amount across platoons in the same company. Isolates from different companies were only found in the largest sub-group (1-1), with as few as 6 core SNPs separating them (including repeat isolates per trainee, the median core SNP distance was 71 across companies, IQR 35-100). Given these companies were separated in time and space, this suggests undetected transmission via an intermediate source.

**Figure 5.**
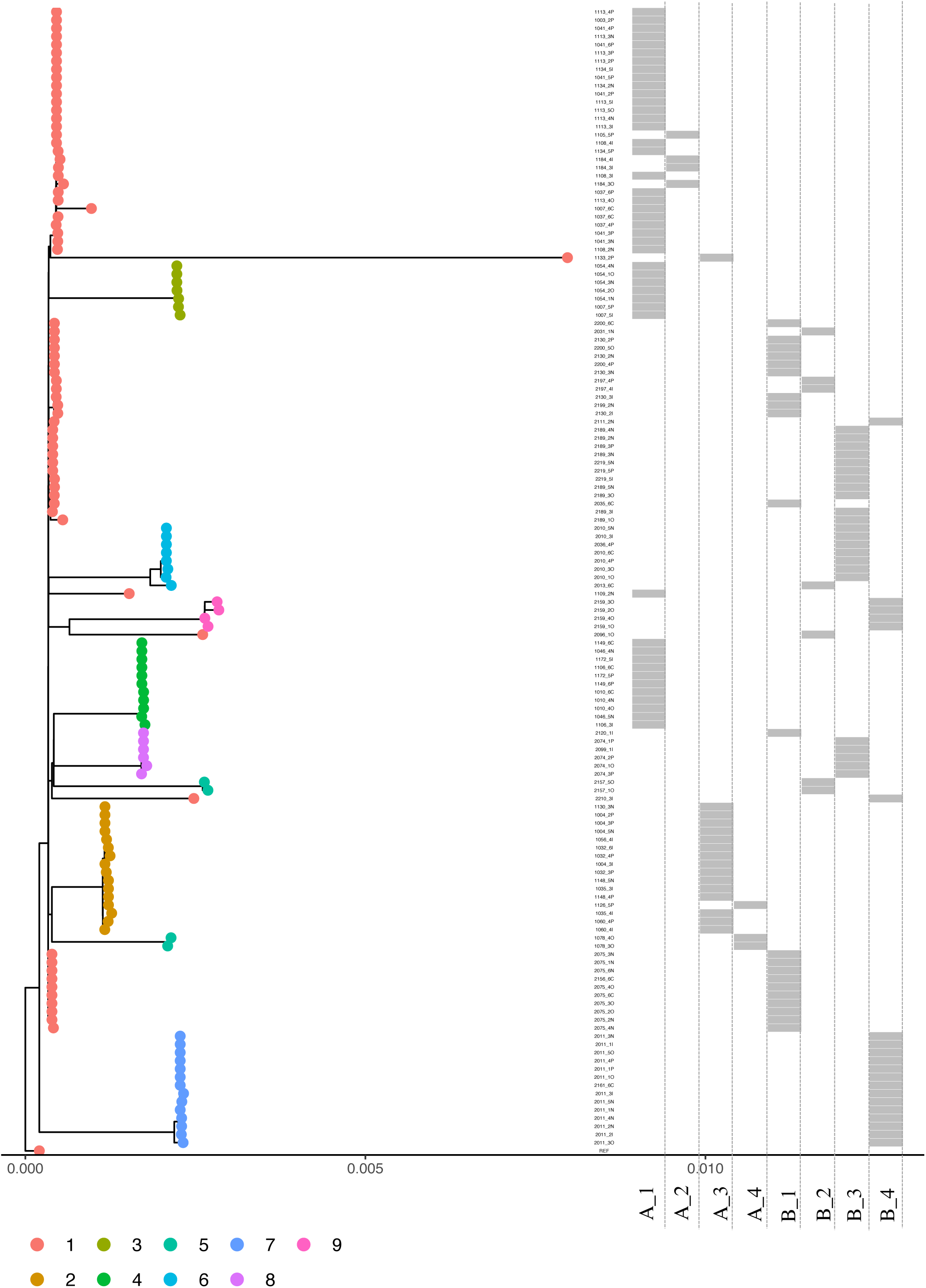
In-depth analysis of Cluster-1 and potential transmission within and across platoons. There were 139 isolates in Cluster-1, as classified by hierBAPs [16, 29] in the overall analysis (Figure 3), 136 of which were ST8. A concatenated core SNP alignment was produced with these isolates. A maximum likelihood tree was produced using IQ-Tree [14] (v1.6.8) and visualized using ggtree [31] in R (v.3.5.2). hierBAPS was again used to identify sub-groups of closely-related isolates within this Cluster, which are indicated in colour. Note that this analysis includes repeat isolates from all trainees; sample names are indicated using the format *Participant Identifier + Visit Number + Anatomical Site of Isolation* (where N=nasal, I=Inguinal, P=Perianal and O=Oropharyngeal), such that repeat samples can be identified. The heatmap indicates epidemiological contact between trainees; the letters (A and B) represent the company and the numbers represent the specific platoon within that company.

Nearly all transmission appears to be within rather than between platoons for Clusters 2 and 4 as well (**Supplementary Figures 2** and **3**). The exception to this was sub-group 1 in Cluster 4, where multiple platoons are implicated; however, all six trainees in this sub-group were colonized at baseline (**Supplementary Results** and **Supplementary Table 6**). There was no evidence of transmission across companies in either Cluster.

### Anatomical sites and their role in transmission

The nares are commonly thought to play a key role in transmission, but > 50% of trainees (regardless of Cluster/ST) were positive only at other anatomical sites (**Table 3**). To examine the impact of this, we repeated our transmission analyses only including isolates from the nares and SSTI (**Supplementary Table 7**) and found that transmission was substantially under-estimated, with as many as 100% of trainees being missed in some Clusters/sub-groups.

### ST8s in context of previous analyses

We compared our contemporaneous samples to historical clinical and colonization isolates from trainees with ST8-associated SSTI at Fort Benning [4, 10] (described further in the **Supplementary Results**), and found historical isolates largely clustered independently of more recent samples (**Supplementary Figure 4**). The median core pairwise distance between these isolates and our 2015 samples was 111 SNPs (IQR 80-137). 99% of pairs (14,187/14,385) were separated by > 15 SNPs, suggesting a single reservoir of MRSA is unlikely to be driving ongoing transmission of ST8-MRSA on base.

## Discussion

MRSA SSTI continues to be a major source of morbidity in military trainees in the United States [2, 3] and other countries [21, 22]. In this study, we integrated genomics with comprehensive epidemiologic data to investigate MRSA transmission in US Army Infantry trainees at Fort Benning, GA, a group known to be at increased risk for MRSA colonization and infection.

We followed individuals prospectively through training for MRSA colonization and SSTI, including all positive colonization isolates from these trainees over time. We found there were multiple STs present at baseline, with subsequent transmission predominantly within, rather than between platoons - reinforcing the importance of close proximity for MRSA transmission.

Consistent with previous ST8 analyses [4, 10], there appeared to be little transmission across training classes (i.e., companies) over time, suggesting no single endemic strain in this context. By contrast, the persistent burden of MRSA colonization and infection is likely due to the serial importation of CA-MRSA strains with each incoming group of new trainees. With > 30,000 trainees from multiple US states and territories cycling through Fort Benning annually, the diversity of MRSA strains being introduced into this single geographic site is likely considerable.

Surprisingly, a large proportion of trainees (27/343, [7.9%]) were positive at baseline or positive shortly after training onset. This suggests that particular phases of training may serve as critical ‘control points’ wherein interventions to reduce transmission and/or SSTI could be implemented. As no specific risk factors for MRSA colonization were identified, we propose that risk of acquisition may be predominantly mediated by exposure to circulating strains and shared training activities, rather than individual hygiene practices - though it is possible the small numbers may preclude detection of significant differences between our comparison groups.

The nares are considered the primary reservoir for *S. aureus* [23]. In the current study, however, we found that more than half (55.3%) of those colonized were exclusively positive at other anatomical sites. Screening of these different sites is therefore critical to accurately detect colonization and resolve transmission. This finding may have important implications for the control of MRSA in military trainees, as MRSA prevalence, transmission, and success of decolonization efforts, have previously been largely estimated from nasal carriage alone [24]. A randomized controlled trial among Army combat medics in training [6] found that, while intranasal mupirocin reduced nasal colonization with MRSA, SSTI rates remained unaffected. Similar results were found in a large-scale, field-based evaluation of chlorhexidine-based body wash [7, 8]. One possibility is that inoculation via fomites may contribute to SSTI, however, another possibility is that trainees colonized exclusively at non-nasal sites (not included in these studies due to the design) served as a constant reservoir for re-colonization/infection. These explanations need not be mutually exclusive. This suggests that, given the negligible levels of resistance to either treatment, mupirocin coupled with chlorhexidine gluconate washes may yet be viable tools for SSTI reduction in this population, potentially coupled with oral antimicrobials [25]. However, for such interventions to work, this would likely require *all* colonized individuals are identified and treated, regardless of anatomical site.

This study has a number of major strengths, including a high participation rate with > 85% of participants completing all visits. Because this was conducted in a military training setting, the schedule, location, type of activities, and risk of MRSA acquisition were relatively uniform for trainees across all platoons at all visits. Additionally, the prospective design, with screening for colonization and collection of corresponding epidemiological data at regular intervals throughout training allowed us to detect such events as they occurred and assess the potential association with specific behaviours that preceded them. This degree of follow-up is rarely feasible outside of a hospital setting. Another strength was the screening of multiple anatomical sites over time; this not only allowed us to more accurately detect colonization than previous studies in this population [6, 7, 24] but examine the role they play in transmission. Finally, previous studies focused exclusively on ST8, with sampling exclusively from cases of SSTI and select controls [4, 10]; by including all STs, we were able to provide the first fully comprehensive analysis of population structure and transmission dynamics of MRSA circulating in this setting.

There are several limitations of this work. Due to resource requirements, we focused recruitment on two (out of 37) training classes from Jun. - Dec. 2015. Though our analysis suggests limited transmission across companies (i.e., classes), it is possible some transmission events were not detected. Permanent staff on base (e.g. drill sergeants, health care personnel, etc.) may also represent a reservoir of MRSA, though our analysis suggest this is less likely (as there were multiple circulating strains, with evidence of importation from outside the base). Staff may also be source of unrecognized transmission, potentially even during in-processing, but unfortunately, were not included in the current study. Similarly, sampling of high touch common surfaces (e.g. countertops, weight benches, wrestling mats, etc.) was not conducted as part of this investigation. Future studies should address these gaps, to better understand the role these may play in MRSA transmission dynamics.

We also note that the included cohorts were purposely chosen based on peak MRSA SSTI seasonality [8], with some studies reporting increased colonization in warmer months [26, 27]. MRSA prevalence in our classes may therefore be higher than would be found in other cohorts. However, baseline prevalence of MRSA colonization among combat medic trainees [6] in Jan. – Dec. 2005 was only 3.9% (based on nasal carriage) and restricting our estimates to nasal carriage yielded a similar proportion (2.0%, p=0.08), suggesting our findings are likely generalizable to other training courses and/or periods. Another potential limitation of this work is that, for privacy reasons, perianal and inguinal swabs were collected by the trainees themselves, rather than by study personnel. Though they were instructed beforehand, it is possible these swabs were not collected in a consistent fashion and may be more likely to yield false negative results. However, we did not find a significant difference in frequency of positivity across anatomical sites. Finally, personal hygiene practices were self-reported at routine study visits for the preceding month, potentially leading to misclassification due to poor recall. However, as colonization status was not known at the time of collecting this information (and was not reported to trainees during the study), we would not expect this misclassification to be differential.

Through this in-depth study, we identified critical timepoints in military training associated with MRSA colonization and highlight the importance of different anatomical sites in mediating this transmission. Given our historical reliance on nasal screening for detection of MRSA colonization, and subsequent interventions to prevent infection, this work may have important implications not only for control of MRSA in the military, but in-patient settings as well. In order to halt MRSA transmission in the military, and other vulnerable populations, increased screening to include these other anatomical sites may be necessary; however, cost and feasibility of such interventions must also be investigated.

## Data Availability

MRSA sequence data is being posted on NCBI’s Sequence Read Archive under BioProject PRJNA587530.

## Acknowledgements

We are thankful for the participation of the US Army Infantry Trainees in our study. We are indebted to the study team of clinical research coordinators, laboratory personnel, and data management staff for their dedication to the project. High-performance computing was done using the Odyssey cluster from the Faculty of Arts and Science, Harvard University. RSL, EVM, AC, CEE, EE, MWE, JWB and WPH have no conflict to declare.

## Contributions

EVM, MWE, and JB designed and ran the parent longitudinal cohort study. WPH and RSL conceived the current genomic study. RSL designed and was responsible for the bioinformatics and epidemiological analyses, designing/preparing the figures and tables, and writing the manuscript. CEE, EE, and AC were responsible for isolating the bacteria, and culture, and AC prepared the sequencing libraries. WPH reviewed the initial draft. All authors were involved in interpreting the data, and reviewed and provided feedback on the manuscript. RSL takes full responsibility for the content of the paper and decision to submit for publication. Please note, the views expressed are those of the authors and do not necessarily reflect the official policy or position of the Henry M. Jackson Foundation for the Advancement of Military Medicine, the Uniformed Services University, the Department of the Army, the Department of Defense, nor the U.S. Government. Mention of trade names, commercial products, or organizations does not imply endorsement by the U.S. Government.

